# Prognostic Significance of Ventilatory Efficiency in Hypersensitivity Pneumonitis

**DOI:** 10.1101/2025.10.06.25337437

**Authors:** Worawat Chumpagern, Paolo Spagnolo, Pailin Ratanawatkul, Michael P. Mohning, Evans R. Fernández Pérez

## Abstract

**Background:** Ventilatory efficiency, measured by a cardiopulmonary exercise testing, is an important metric for evaluating cardiopulmonary diseases. However, its relationship with clinical outcomes in hypersensitivity pneumonitis remains unclear.

**Objective:** To determine if ventilatory efficiency correlates with various stages of severity and predicts mortality in HP.

**Study Design and Methods:** We conducted a retrospective cohort study from 2009 to 2019 involving patients with non-fibrotic (nfHP) and fibrotic HP (fHP), and idiopathic pulmonary fibrosis (IPF), which served as the comparison group. CPET variables, including ventilatory efficiency (slope and intercept from the linear regression model of ventilation (VE) vs. carbon dioxide output (VCO2) and the nadir ratio), were assessed across various forced vital capacity (FVC%) and diffusing capacity for carbon monoxide (DLCO%) ranges. We used a multivariate logistic regression model to identify predictors of five-year mortality and Kaplan-Meier plots to assess survival.

**Results:** 164 patients were analyzed, with a mean age of 65.6±12.0 years and 54.3% male. Twenty-five (15.2%) had nfHP, 66 (40.2%) fHP, and 73 (44.5%) IPF. Overall, an increase in the VE/VCO2 slope and nadir was observed as the DLCO% decreased, but there was no significant change with a decrease in FVC%, while the intercept remained unchanged. Patients with pulmonary hypertension (35%, 57/164) and fHP or IPF had significantly elevated VE/VCO2 slope and nadir when compared to patients without pulmonary hypertension or nfHP, respectively. A VE/VCO2 slope ≥42 (AUC 0.67; 95% CI 0.53-0.82) was identified as a predictor for survival (HR 3.65; 95% CI, 1.04-12.80). Restricting the analysis to patients with HP showed similar results (HR 5.49; 95% CI, 1.19 – 33.54).

**Interpretation:** In patients with HP, VE/VCO2 was associated with the presence of pulmonary fibrosis, PH, reduced DLCO, and a higher mortality risk. A VE/VCO2 slope threshold of 42 may be a useful prognostic marker for stratifying patients into low-risk and high-risk groups.

## Background

Hypersensitivity pneumonitis (HP) and idiopathic pulmonary fibrosis (IPF) represent two of the most prevalent forms of interstitial lung disease (ILD) encountered in pulmonary clinics globally. A subset of patients diagnosed with HP may progress to develop pulmonary fibrosis.^1,2^ Similar to patients with IPF, individuals with fibrotic and nonfibrotic HP commonly experience dyspnea. While dyspnea is a primary symptom for both IPF and HP, accompanying comorbidities, such as pulmonary hypertension (PH), can exacerbate this symptom, further limiting physical activity and significantly diminishing quality of life and independence over time.

Cardiopulmonary exercise testing (CPET) provides valuable insights into functional capacity, cardiovascular function, ventilatory efficiency, and pulmonary gas exchange in patients with ILD. Among the key metrics evaluated, ventilatory efficiency—measured as the ratio of minute ventilation to carbon dioxide production (VE/VCO2)—is frequently compromised in individuals with ILD. This impairment indicates that patients must engage in higher levels of ventilation to expel the same quantity of carbon dioxide. Consequently, this inefficiency contributes to the experience of breathlessness and limits exercise capacity.

To date, there is only one study that examines the CPET characteristics in 28 patients with HP compared to 13 control subjects.^3^ Nonetheless, the role of ventilatory inefficiency in the risk assessment and prognostic evaluation of ILD has been limited to IPF.^4^ Furthermore, there exists a significant gap in the literature regarding the relationships among ventilatory efficiency, lung function impairment, and pulmonary hypertension in individuals presenting for initial evaluation diagnosed with HP with and without pulmonary fibrosis.

Hence, we aimed to investigate patients with and without fibrotic HP, and IPF as a comparison group: (1) the relationship between ventilatory inefficiency, pulmonary fibrosis, and PH, stratified by forced vital capacity (FVC) and diffusing capacity for carbon monoxide (DLCO); and (2) the relationship of ventilatory inefficiency and survival.

## Study Design and Methods

### Population

This institutional review board-approved (HS3248) retrospective cohort study was conducted at National Jewish Health (NJH), a multispecialty tertiary referral center. While the main goal was to evaluate CPET ventilatory efficiency in HP, cases of IPF were analyzed as disease comparison group. The NJH ILD research database was queried for consecutive adult patients referred for evaluation with HP and IPF who had a maximum cardiopulmonary test between 2009 and 2019 as part of their initial evaluation. The diagnosis of HP and IPF was determined through a structured multidisciplinary discussion that utilized all available data at the time of the initial assessment in the ILD clinic, which served as the baseline data for all analyses.

The presence of pulmonary fibrosis was defined by the presence of reticular abnormality and/or, traction bronchiectasis and/or, architectural distortion, and/or honeycombing on HRCT at time of diagnosis.

For baseline, pulmonary hypertension assesments, PFTs, and CPET performed within 30 days of diagnosis were retrived from the research database:

a. The diagnosis of pulmonary hypertension is based on right-heart cardiac catheterization (mean pulmonary artery pressure (mPAP) ≥ 25 mm and mean pulmonary wedge pressure (mPCWP) ≤ 15 mm) or a transthoracic echocardiogram (right ventricular systolic pressure (RVSP) ≥ 35 mmHg or peak tricuspid regurgitant velocity (TRV) ≥ 2.8 m/s).^5^ Patients with post-capillary pulmonary hypertension were excluded.
b. PFT including spirometry, maximum voluntary ventilation (MVV), and diffusion capacity, were conducted following the guidelines set by the American Thoracic Society and the European Respiratory Society.^6,7^ The reference equation is outlined in earlier publications.^8^ All indices from the pulmonary function tests (FVC and DLCO% predicted) were categorized into four stages based on z-scores: preserved: >-2.6, mild: - 1.65 to -2.5, moderate: -2.51 to -4.0, and severe: <-4.1.^9^
c. Every patient underwent room-air maximal cardiopulmonary exercise testing using a bicycle ergometer at our facility, following the same standardized protocol. A radial arterial line was placed before the test. The ramp protocol was endorsed, consisting of three minutes of pedaling at zero workload and an incremental workload range of 5 to 40 watts, while maintaining a pedal speed of at least 60 rpm, until the test termination criteria were met. Then, the recovery phase was initiated. Physiological data were collected. Oxygen uptake (VO_2_ , STPD), carbon dioxide production (VCO_2_ , STPD), ventilation (VE, BTPS), tidal volume, respiratory frequency (fR) and end-tidal CO2 partial pressure (PETCO_2_ ) were measured breath-by-breath (Ultima Cardio2 System, Minnesota, USA). Arterial oxygen saturation was monitored using a pulse oximeter (SpO2). The anaerobic threshold (AT) was identified by the V-slope method along with supporting ventilatory and pulmonary gas exchange criteria. The physiological V_D_/V_T_ was calculated by using PETCO_2_ and PaCO_2_ data from arterial blood gas. Ventilatory efficiency was assessed using breath-to-breath data from each patient.^10^ A linear regression model identifies the slope and y-intercept between V′CO_2_ (x-axis) and V′E (y-axis), omitting data that exceeds the ventilatory compensatory point. The nadir V′E/V′CO_2_ was calculated by averaging the three lowest consecutive 0.5-minute data points.

### Statistical analysis

Descriptive statistics of baseline and longitudinal data were generated. The normality of frequency distributions was assessed with the Shapiro-Wilk test. Values are presented as means ± standard deviation (SD), unless stated otherwise. A p-value of less than 0.05 was considered statistically significant for all analyses. To evaluate differences between proportions, a chi-square test for homogeneity was employed. Differences between means were analyzed using either the unpaired t-test or the Mann-Whitney U test, depending on the distribution of the data. The ANOVA with Tukey’s method was utilized for comparisons across varying degrees of FVC and DLCO% predicted ranges.

Receiver operating characteristic curve (AUC) analysis was conducted to assess the prognostic significance of the variables, identifying optimal cut-off points by using Youden’s method. AUCs are reported along with 95% confidence intervals. Kaplan–Meier survival curves were generated, and the log-rank test was used to compare survival times between groups. Vital status was ascertained on December 31, 2024.

Hazard ratios and their 95% confidence intervals were calculated using Cox proportional hazards analysis. Also, correlation analysis was performed to address multicollinearity.

Predictors were included in the model if their relationships with 5-year mortality met a significance threshold of P < 0.10. After establishing a full model that included all candidate predictors, we applied bidirectional elimination to derive a reduced model. The final multivariate logistic regression model comprised independent predictors of 5-year mortality, with the criterion for retaining covariates set at P < 0.05.

All analyses were conducted using R Studio version 2024.12.1+563.

## Results

### Baseline characteristics

The study flowchart is shown in **Figure 1**. From a total of 659 patients with ILD and CPET, 164 had a multidisciplinary HP or IPF diagnosis and were included in the final analysis. The mean age was 65.6 ± 12.0 years, and 54.3% were male. Twenty-five (15.2%) were diagnosed with non-fibrotic HP, 66 (40.2%) with fibrotic HP, and 73 (44.5%) with IPF.

**Figure 1.**
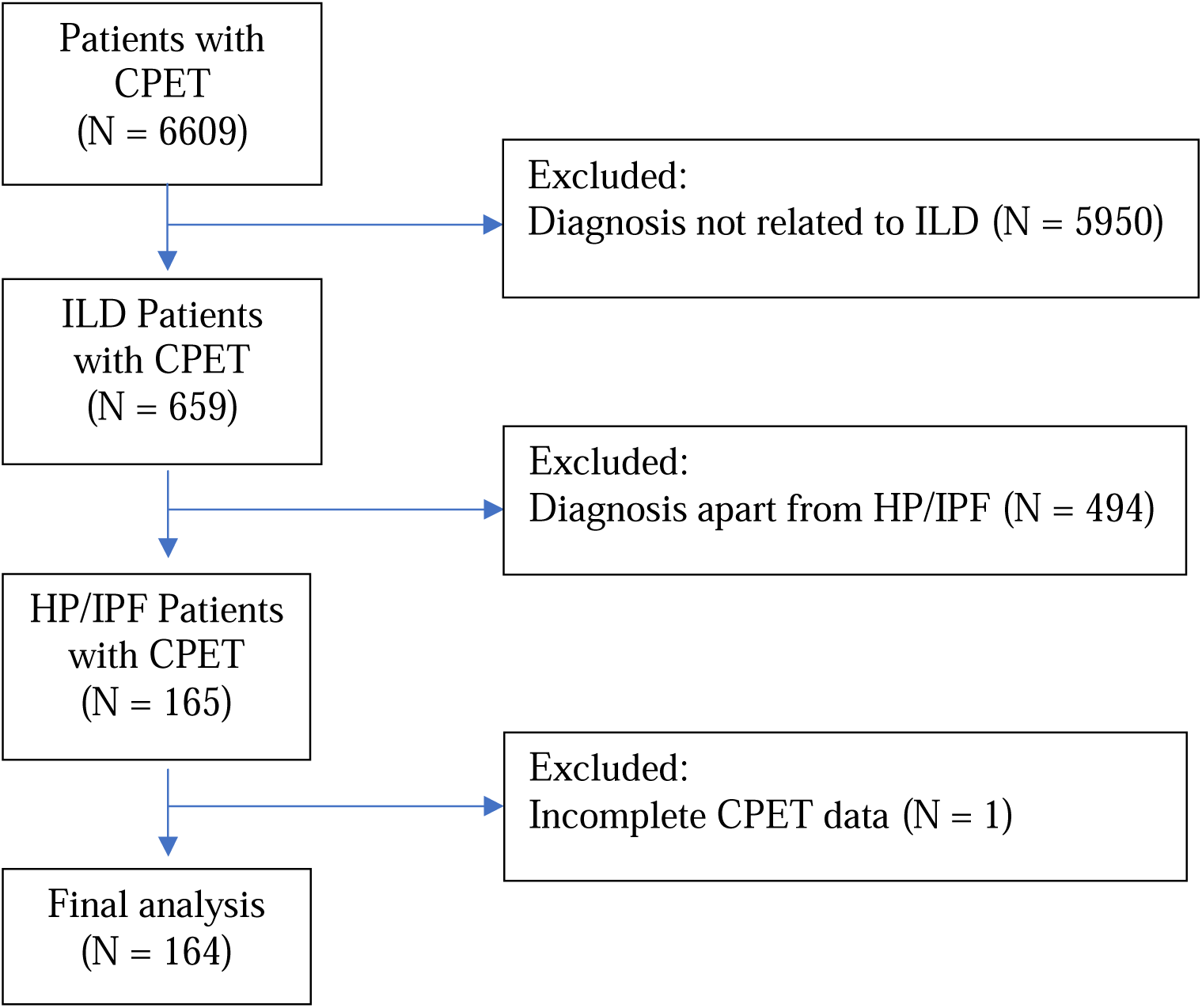
Flowchart of the study.

The baseline characteristics, lung function tests and CPET variables stratified by non-fibrotic HP, fibrotic HP and IPF are shown in **Table 1**. Compared to non-fibrotic HP, fibrotic HP and IPF patients were generally older, more often male, had more often pulmonary hypertension and displayed more restrictive lung function (lower FVC%, DLCO% and TLC%). All groups had similar FEV1/FVC% and RV/TLC% ratios.

**Table 1.** Baseline Clinical Characteristics.

| Characteristics | Non-fibrotic HP<br>(n = 25) | Fibrotic HP<br>(n = 66) | IPF<br>(n = 73) | P<br>Value |
| --- | --- | --- | --- | --- |
| Age, y | 57.8 ± 13.6 | 60.6 ± 11.2 | 72.9 ± 7.7<sup>a,b</sup> | **<0.001** |
| Male | 7 (28.0) | 27 (40.9) | 55 (75.3)<sup>a,b</sup> | **<0.001** |
| BMI, kg/m<sup>2</sup> | 29.5 ± 5.2 | 29.8 ± 5.9 | 28.2 ± 4.5 | 0.172 |
| Former smoker | 0 (0%) | 1 (1.5%) | 4 (5.5%) | 0.64 |
| Pulmonary hypertension | 4 (16.0%) | 22 (33.3%) | 31 (42.5%)<sup>a</sup> | **0.05** |
| Pulmonary function tests |  |  |  |  |
| FVC predicted, % | 84.4 ± 14.1 | 70.8 ± 15.2<sup>a</sup> | 79.1 ± 20.4<sup>b</sup> | **<0.001** |
| FEV<sub>1</sub> predicted, % | 87.2 ± 13.6 | 74.3 ± 16.3<sup>a</sup> | 86.4 ± 20.9<sup>b</sup> | **<0.001** |
| FEV<sub>1</sub>/FVC | 81.6 ± 5.8 | 82.5 ± 8.3 | 83.3 ± 6.7 | 0.58 |
| TLC predicted, % | 98.1 ± 34.0 | 76.9 ± 14.0<sup>a</sup> | 73.7 ± 14.9<sup>a</sup> | **<0.001** |
| RV predicted, % | 114.2 ± 32.0 | 93.8 ± 24.0<sup>a</sup> | 74.1 ± 18.4<sup>a,b</sup> | **<0.001** |
| RV/TLC predicted % | 94.7 ± 48.9 | 110.2 ± 43.0 | 101.4 ± 19.7 | 0.210 |
| DLCO predicted, % | 88.0 ± 19.8 | 75.1 ± 19.8<sup>a</sup> | 69.8 ± 19.8<sup>a</sup> | **<0.001** |
| Cardiopulmonary exercise test parameters |  |  |  |  |
| Work, Watt | 124.2 ± 55.6 | 102.4 ± 41.8<sup>a</sup> | 104.5 ± 37.2 | 0.07 |
| Work %predicted, % | 92.7 ± 28.8 | 78.5 ± 34.8<sup>a</sup> | 70.2 ± 23.7<sup>a</sup> | **<0.001** |
| VO<sub>2</sub> Max, ml/kg/min | 18.8 ± 6.2 | 16.1 ± 5.3<sup>a</sup> | 16.3 ± 4.1 | **0.05** |
| VO<sub>2</sub> Max predicted, % | 92.8 ± 20.5 | 84.2 ± 26.6 | 80.0 ± 25.5 | 0.09 |
| ΔVO<sub>2</sub>/ΔWR, ml/min/watt | 9.69 ± 1.49 | 9.18 ± 1.58 | 9.08 ± 1.91 | 0.30 |
| AT/Predicted VO<sub>2</sub>, % | 51.3 ± 14.1 | 46.9 ± 14.4 | 49.9 ± 12.5 | 0.26 |
| VE max, L/min | 72.2 ± 22.0 | 61.2 ± 21.5 | 71.1 ± 21.7<sup>b</sup> | **0.01** |
| VE max/MVV, % | 73.0 ± 13.6 | 75.2 ± 22.0 | 75.3 ± 17.6 | 0.86 |
| Breathing reserve, L/min | 29.0 (12.7, 43.6) | 18.3 (8.07, 33.3) | 23.3 (11.5, 37.1) | 0.44 |
| V<sub>T</sub> max, L | 1.80 ± 0.61 | 1.42 ± 0.46<sup>a</sup> | 1.66 ± 0.51<sup>b</sup> | **0.002** |
| fR max, rpm | 41.1 ± 7.8 | 44.2 ± 11.5 | 45.2 ± 11.9 | 0.29 |
| HR max/HR predicted, % | 88.7 ± 8.8 | 87.9 ± 13.3 | 85.8 ± 13.2 | 0.50 |
| Oxygen pulse predicted,% | 105.2 ± 23.3 | 96.2 ± 30.6 | 92.5 ± 27.0 | 0.16 |
| Borg Dyspnea | 8.36 ± 1.66 | 7.86 ± 1.91 | 8.32 ± 1.62 | 0.23 |
| SpO<sub>2</sub> at rest, % | 94.5 ± 2.7 | 94.1 ± 2.7 | 92.5 ± 3.1<sup>a,b</sup> | **0.001** |
| SpO<sub>2</sub> at peak exercise, % | 90.1 ± 6.2 | 87.1 ± 6.1<sup>a</sup> | 85.1 ± 6.2<sup>a</sup> | **0.002** |
| PETCO<sub>2</sub> at rest, mmHg | 30.5 ± 8.1 | 31.6 ± 3.7 | 30.0 ± 3.4 | 0.12 |
| PETCO<sub>2</sub> at peak exercise, mmHg | 29.6 ± 4.8 | 32.1 ± 4.6<sup>a</sup> | 28.6 ± 4.5<sup>b</sup> | **<0.001** |
| V<sub>D</sub>/V<sub>T</sub> at rest | 0.29 ± 0.08 | 0.36 ± 0.07<sup>a</sup> | 0.41 ± 0.07<sup>a,b</sup> | **<0.001** |
| V<sub>D</sub>/V<sub>T</sub> at peak exercise | 0.24 ± 0.09 | 0.29 ± 0.08<sup>a</sup> | 0.36 ± 0.09<sup>a</sup> | **<0.001** |
| VE-VCO<sub>2</sub> Slope | 34.2 ± 9.5 | 35.3 ± 7.1 | 42.0 ± 9.9<sup>a,b</sup> | **<0.001** |
| VE-VCO<sub>2</sub> Slope ≥ 42 | 3 (12.0%) | 13 (19.7%) | 32 (43.8%)<sup>a,b</sup> | **<0.001** |
| Nadir VE/VCO<sub>2</sub> | 33.0 ± 6.4 | 35.2 ± 6.3 | 38.5 ± 7.5<sup>a,b</sup> | **<0.001** |
| Nadir V'E/V'CO<sub>2</sub> ≥ 36.5 | 5 (20%) | 21 (31.8%) | 36 (49.3%)<sup>a,b</sup> | **0.01** |
| Values are No. (%), mean ± SD, median (IQR) or as otherwise indicated. AT = anaerobic threshold; BMI = body mass index; DLCO = diffusing capacity for carbon monoxide; FEV<sub>1</sub> = forced expiratory volume in 1 minute; fR = respiratory frequency; FVC = forced vital capacity; HP = hypersensitivity pneumonitis; HR = heart rate; PETCO<sub>2</sub> = end-tidal CO<sub>2</sub> partial pressure; RV = residual volume; SpO<sub>2</sub> = pulse oximeter; TLC = total lung capacity; V<sub>D</sub>/V<sub>T</sub> = dead space fraction; VE = minute ventilation; VCO<sub>2</sub> = carbon dioxide production; VO<sub>2</sub> = oxygen consumption; V<sub>T</sub> = tidal volume; WR = work rate |  |  |  |  |
| <sup>a</sup> Significantly different from Non-fibrotic HP |  |  |  |  |
| <sup>b</sup> Significantly different from Fibrotic HP |  |  |  |  |

On CPET, compared to non-fibrotic HP, fibrotic HP and IPF patients had lower predicted maximum workload, maximum VO_2_, higher end-tidal carbon dioxide partial pressure at peak exercise, lower ventilatory threshold, lower oxygen saturation at peak exercise, and increased deadspace fraction (**Table 1, e-Table 1**).

### Ventilatory efficiency is more closely associated with gas exchange abnormalities than restrictive impairments

Overall, ventilatory efficiency remained constant across different levels of decreased FVC% Z-scores (**Figure 2**). However, the VE/VCO2 slope and nadir increased as DLCO% Z-scores decreased, while the VE/VCO2 intercepts remained constant.

**Figure 2:**
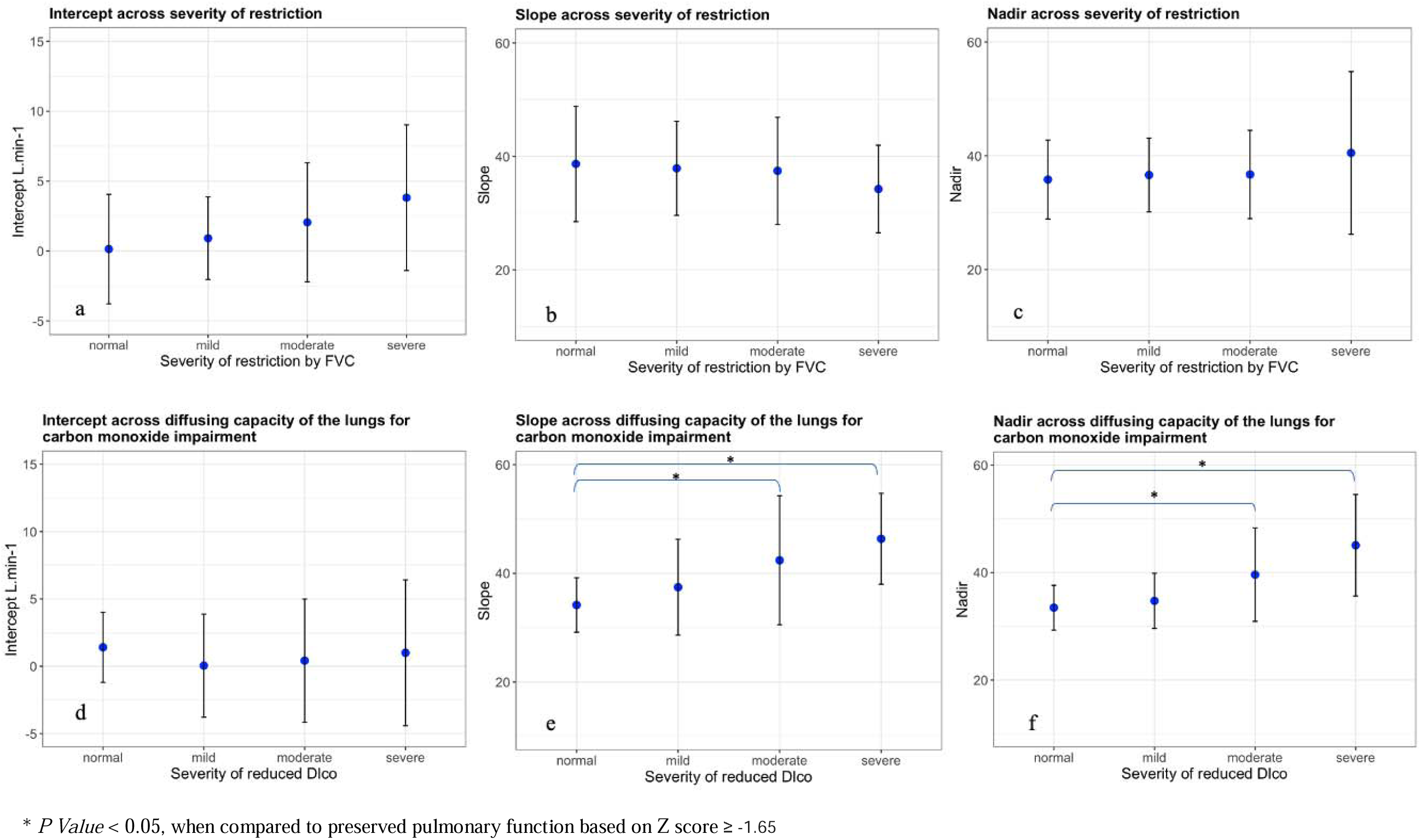
Ventilatory efficiency in relation to various levels of lung function shows that the VE intercept, VE/VCO2 slope, and nadir VE/VCO2 do not significantly vary with FVC decline (a-c). However, both VE/VCO2 slope and nadir VE/VCO2 show an inverse correlation with reduced DLCO (e,f), while the V′E intercept remains consistent throughout (d).

Significant correlation was found between the VE/VCO2 slope and nadir with the DLCO Z-score (VE/VCO2 slope vs. DLCO Z-score: r = -0.42 (*P* = <0.001), VE/VCO2 nadir vs. DLCO Z-score: r = -0.51, *P* = <0.001). Also, maximum workload, maximum VO2, SpO2, and VD/VT at maximum exercise were correlated with decreasing DLCO (**e-Table 2**, **e-Table 3**).

### Ventilatory efficiency is associated to the presence of pulmonary fibrosis and pulmonary hypertension

Compared to non-fibrotic HP (n=25), fibrotic HP and IPF patients (n=139) had an increased VE/VCO2 slope and nadir with a relatively constant intercept (**Figure 3d-f**, **e-Table 1**). Restricting the analysis to patients with HP showed similar results (**Table 1**). Likewise, patients with PH (n=57) also showed elevated VE/VCO2 slope and nadir, in contrast to those without PH (n=107). However, patients with PH experienced a decreased oxygen pulse during exercise (**e-Table 4**).

**Figure 3.**
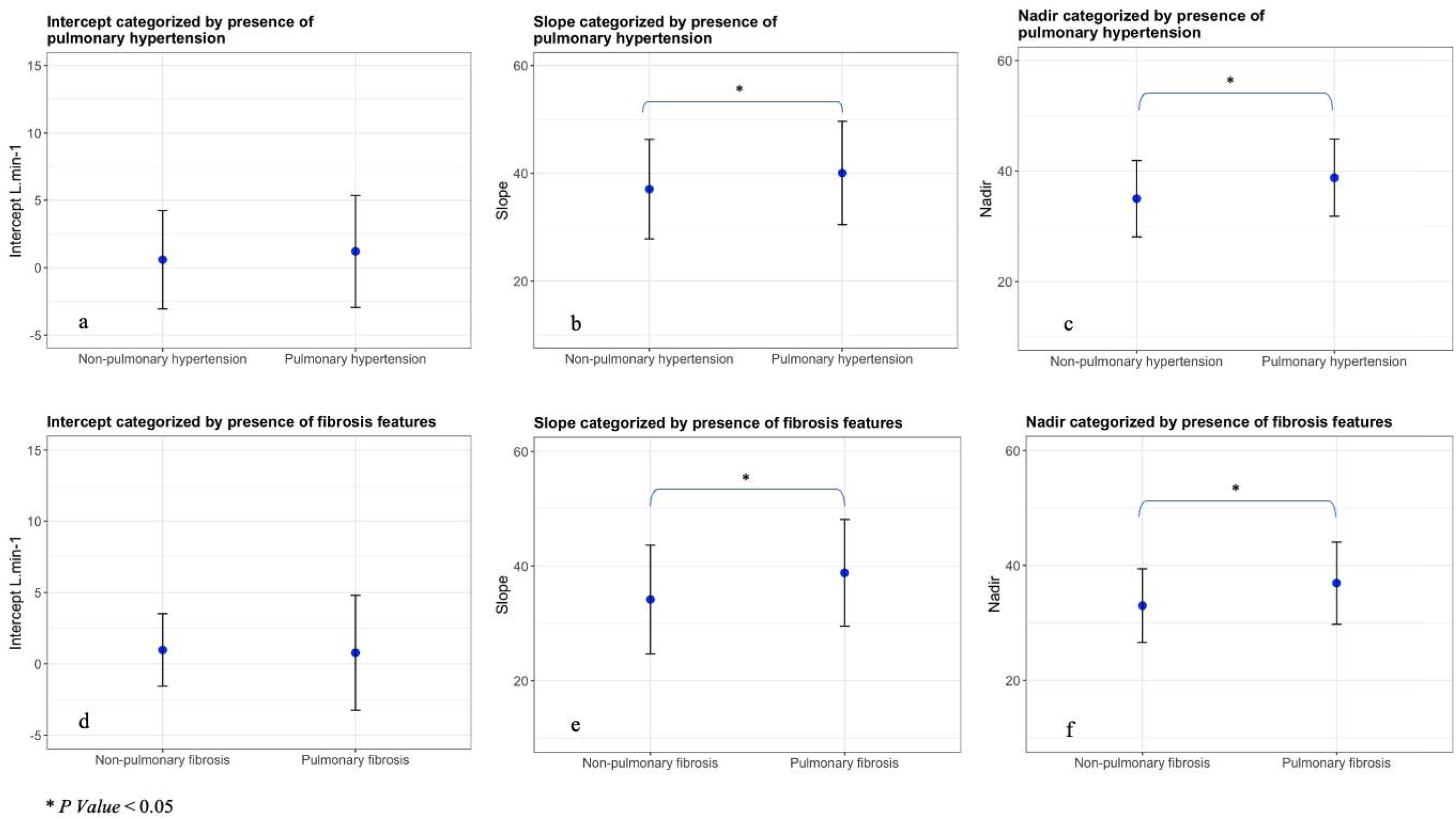
Relationship between ventilatory efficiency and PH and pulmonary fibrosis. Both the VE/VCO2 slope and nadir increased in the presence of PH (b, c) and pulmonary fibrosis (e, f). The VE intercept remains unchanged, regardless of the presence of PH (a), or pulmonary fibrosis (d).

### Ventilatory efficiency predict survival

**Table 2** shows the key predictors of 5-year mortality, as determined through both univariable and multivariable Cox proportional hazards models. The 1-minute heart rate recovery, VE/VCO2 slope and VE/VCO2 nadir were significant predictors. VE/VCO2 nadir was excluded from the multivariate analysis due to collinearity with VE/VCO2 slope.

**Table 2.** Univariable and multivariable Cox proportional Hazard Analysis for Baseline Clinical and CPET variables.

| **Univariable regression** | **Hazard ratio** | **P Value** |
| --- | --- | --- |
| Age | 1.06 (1.01 – 1.16) | **0.008** |
| Male | 1.49 (0.52 – 4.19) | 0.44 |
| Pulmonary Hypertension | 1.71 (0.62 – 4.74) | 0.29 |
| FVC, L | 0.63 (0.33 – 1.21) | 0.17 |
| FVC predicted, % | 0.99 (0.96 – 1.01) | 0.51 |
| FEV<sub>1</sub>, L | 0.64 (0.28 – 1.43) | 0.28 |
| FEV<sub>1</sub> predicted, % | 0.99 (0.97 – 1.02) | 0.98 |
| Dlco, ml/min/mmHg | 0.87 (0.77 – 0.98) | **0.02** |
| Dlco %predicted | 0.92 (0.94 – 1.00) | 0.08 |
| TLC predicted, % | 0.97 (0.94 – 1.01) | 0.22 |
| RV predicted, % | 0.98 (0.96 – 1.00) | 0.10 |
| WR | 0.98 (0.96 – 0.99) | **0.01** |
| Maximum VO<sub>2</sub> | 0.90 (0.80 – 1.01) | 0.08 |
| VE max, L/min | 0.98 (0.96 – 1.00) | 0.22 |
| V<sub>T</sub> max, L | 0.45 (0.15 – 1.33) | 0.15 |
| 1-minute heart rate recovery | 0.90 (0.83 – 0.97) | **0.01** |
| 1-minute heart rate recovery ≤ 11 beats | 5.06 (1.45 – 17.7) | **0.01** |
| SpO<sub>2</sub> at rest, % | 1.01 (0.87 – 1.18) | 0.82 |
| SpO<sub>2</sub> at peak exercise, % | 0.98 (0.90 – 1.07) | 0.78 |
| SpO<sub>2</sub> < 89% at peak exercise | 1.73 (0.65 – 4.54) | 0.37 |
| PETCO<sub>2</sub> at maximum exercise | 0.86 (0.77 – 0.95) | **0.003** |
| VE-VCO<sub>2</sub> Slope | 1.08 (1.04 – 1.13) | **<0.001** |
| VE-VCO<sub>2</sub> Slope ≥ 42 | 4.62 (1.76 – 12.11) | **<0.001** |
| Nadir VE/VCO<sub>2</sub> | 1.15 (1.07 – 1.22) | **<0.001** |
| Nadir VE/VCO<sub>2</sub> ≥ 36.5 | 7.07 (2.47 – 20.23) | **<0.001** |
| **Multivariable regression** | **Hazard ratio** | **P Value** |
| VE/VCO<sub>2</sub> Slope ≥ 42 | 3.65 (1.04 – 12.80) | **0.04** |
| 1-minute heart rate recovery ≤ 11 beats | 6.95 (1.41 – 28.80) | **0.01** |
| WR | 0.99 (0.97 – 1.01) | 0.43 |
| Dlco = diffusing capacity for carbon monoxide; FEV<sub>1</sub> = forced expiratory volume in 1 minute; fR = respiratory frequency; FVC = forced vital capacity; HR = heart rate; PETCO<sub>2</sub> = end-tidal CO<sub>2</sub> partial pressure; RV = residual volume; SpO<sub>2</sub> = pulse oximeter; TLC = total lung capacity; VE = minute ventilation; VCO<sub>2</sub> = carbon dioxide production; VO<sub>2</sub> = oxygen consumption; V<sub>T</sub> = tidal volume; WR = work rate |  |  |

The Kaplan-Meier curve depicting the relationship between the VE/VCO2 slope, VE/VCO2 nadir and five-year survival is presented in **Figure 4**. Seventeen (10.3%) died within 5 years (5 with fibrotic HP and 12 with IPF). Median survival was 21 months. A cut-off point for the VE/VCO2 slope that best divided the overall study cohort into high-risk and low-risk groups was determined to be 42, based on an AUC of 0.67, 95% CI 0.53 to 0.82. For the nadir VE/VCO2, the cut-off point was established at 36.5 (AUC of 0.73, 95% CI 0.63 to 0.84).

**Figure 4.**
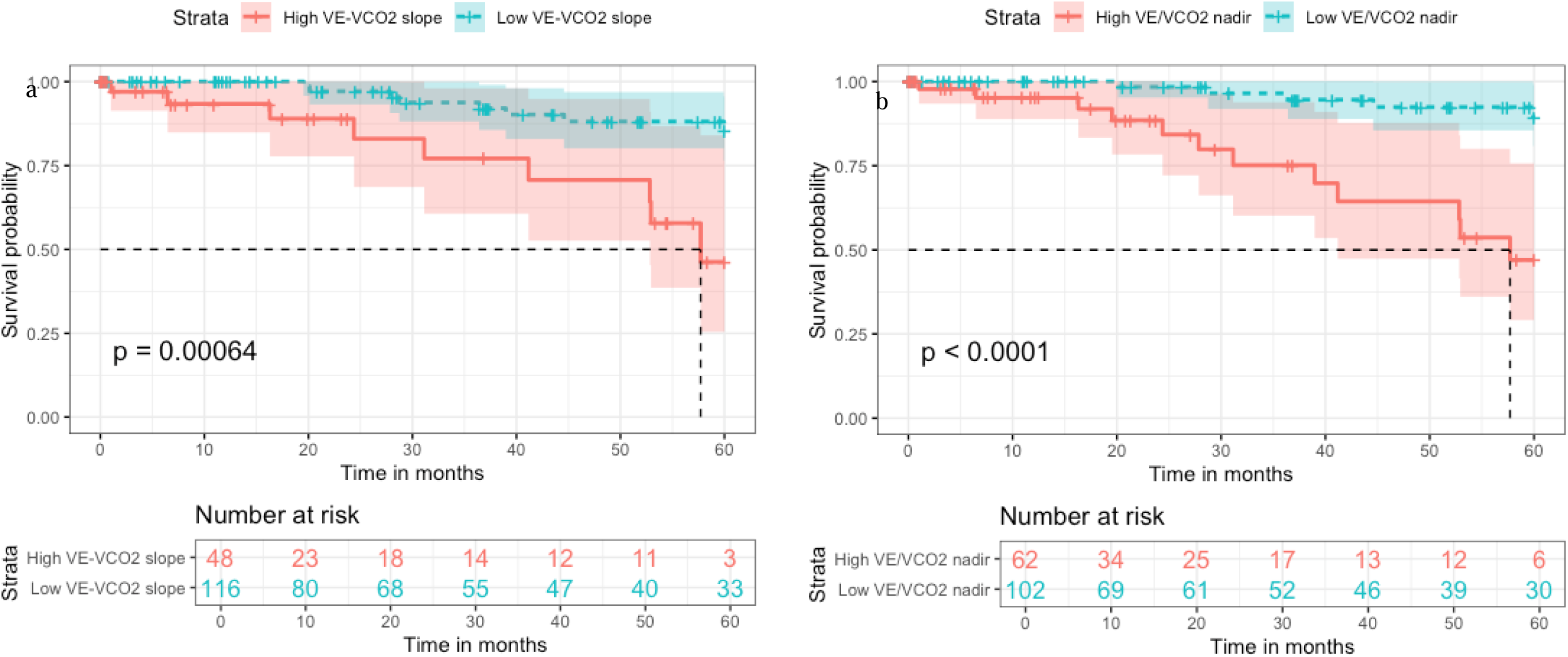
Assessment of Ventilatory Efficiency as a Predictor of 5-Year Survival: The VE/VCO2 slope is categorized as high (≥ 42) or low (< 42) (a), while the nadir VE/VCO2 is classified as high (≥ 36.5) or low (< 36.5) (b).

Restricting the analysis to patients with HP revealed that a VE/VCO2 slope greater than 42 distinguished low-risk and high-risk groups. (HR 5.49; 95% CI, 1.19 to 33.54, **e-Figure 1**).

## Discussion

In a cohort of patients with non-fibrotic HP, fibrotic HP and IPF, we found that exercise ventilatory inefficiency was: (1) significantly associated with the presence of pulmonary fibrosis and PH, (2) worsened significantly with declining DLCO, and (3) identified patients at greater risk of poor outcome. Specifically, we found that a baseline threshold VE/VCO2 slope of ≥42 predicts mortality in these patients.

Although not the main focus of our study, our multivariable regression model indicated that a 1-minute heart rate recovery of ≤ 11 beats is a significant predictor of mortality. Other studies have also shown that this metric correlates with clinical outcomes in cardiopulmonary diseases. ^11,12^ This abnormal response may stem from an impaired autonomic reaction post-exercise, chronotropic incompetence due to right heart dysfunction, and hypoxemia leading to sympathovagal imbalance. Indeed, decreased heart rate recovery was notably more pronounced in patients with PH (**e-Table 4**), as well as in those with fibrotic HP and IPF, compared to those with non-fibrotic HP (**e-Table 1**).

The findings of this study align with earlier research indicating that patients with ILD exhibit an elevated VE/VCO2 slope and nadir. This phenomenon can be attributed to a combination of increased physiological dead space, compensatory tachypnea, and heightened ventilatory drive due to hypoxemia.^13,14^ Indeed, in contrast to non-fibrotic HP, patients with fibrotic HP and IPF, who have greater baseline disease severity, displayed a higher ventilation-perfusion ratio and a lower arterial carbon dioxide tension during maximal exercise (**e-Table 1**). We also observed that the VE/VCO2 intercept was normal and constant in our patient cohort, independent of the presence of pulmonary fibrosis or PH, suggesting there is no excessive ventilation at rest, as the factors described above, particularly increase dead space ventilation, contributing to ventilatory inefficiency in ILD, become more pronounced during exercise. In addition, a significant number of patients in our study (57/164, 35%) had PH, which can further exacerbate ventilatory inefficiency due to an increased V/Q mismatch and blunted cardiac output during exercise.

A key finding of this study is that VE/VCO2 demonstrated a stronger correlation with a progressive decline in DLCO compared to a progressive decline in FVC, likely because both VE/VCO2 and DLCO are sensitive measures of gas exchange across the alveolar-capillary membrane, whereas FVC primarily measures lung volume.

Previous studies, particularly those involving patients with IPF—who were included as a comparison group in our study—have demonstrated that exercise ventilatory inefficiency serves as a pathophysiological marker in various cardiopulmonary diseases. This study represents the largest analysis validating this finding among patients with HP, presenting for initial ILD evaluation, while controlling for the presence of lung fibrosis and PH, which are known to be important predictors of poor outcome.

Baseline lung function has been demonstrated to predict survival in cohorts with HP. Specifically, a lower FVC% and DLCO% are associated with increased mortality.^15–17^ While baseline metrics, such as pulmonary function tests, provide a static measure of lung capacity, evaluating exercise ventilatory efficiency offers a dynamic and sensitive assessment of disease severity and progression, which can enhance the tools available for risk stratification in patients with HP.

The limitations of this study include its retrospective design and that we do not specifically account for post-CPET treatment effects on survival. Furthermore, our cohort lacked information on the specific causes of death. While the prognostic value of ventilatory efficiency at levels ranging from >34 to >46 has been reported to predict survival in IPF, to the best of our knowledge, the prognostic significance of this metric has not been previously investigated in HP. Although this is the first study to provide a practical threshold for VE/VCO that includes patients with HP, the results should be replicated in an independent cohort of patients with HP.

## Interpretation

We found that in a cohort of patients with non-fibrotic HP, fibrotic HP, and IPF undergoing room-air maximal CPET using a bicycle ergometer, ventilatory inefficiency, indicated by an elevated VE/VCO2 slope and nadir, was associated with the presence of pulmonary fibrosis, PH, reduced DLCO, and an increased risk of mortality. Notably, a VE/VCO2 slope threshold of 42 or higher may serve as a useful prognostic marker, helping stratify patients into low-risk and high-risk groups.

## Data Availability

All data produced in the present work are contained in the manuscript

## Author’s contributions

Study conception and design: E.R.F.P. Data collection: W.C., P.S., P.R., E.R.F.P. Data analysis: W.C., P.R., and E.R.F.P. Initial manuscript draft: E.R.F.P. Manuscript revision for critically important intellectual content: all authors.

## Abbreviations list

AT: anaerobic threshold
BMI: body mass index
DLCO: diffusing capacity for carbon monoxide
FEV_1_: forced expiratory volume in 1 minute
fR: respiratory frequency
FVC: forced vital capacity
HP: hypersensitivity pneumonitis
HR: heart rate
PETCO_2_: end-tidal CO_2_ partial pressure
RV: residual volume
SpO_2_: pulse oximeter
TLC: total lung capacity
V_D_/V_T_: dead space fraction
VE: minute ventilation
VCO2: carbon dioxide production
VO_2_: oxygen consumption
V_T_: tidal volume
WR: work rate
ILD: interstitial lung disease.

## Supplement

**e-Table 1.**
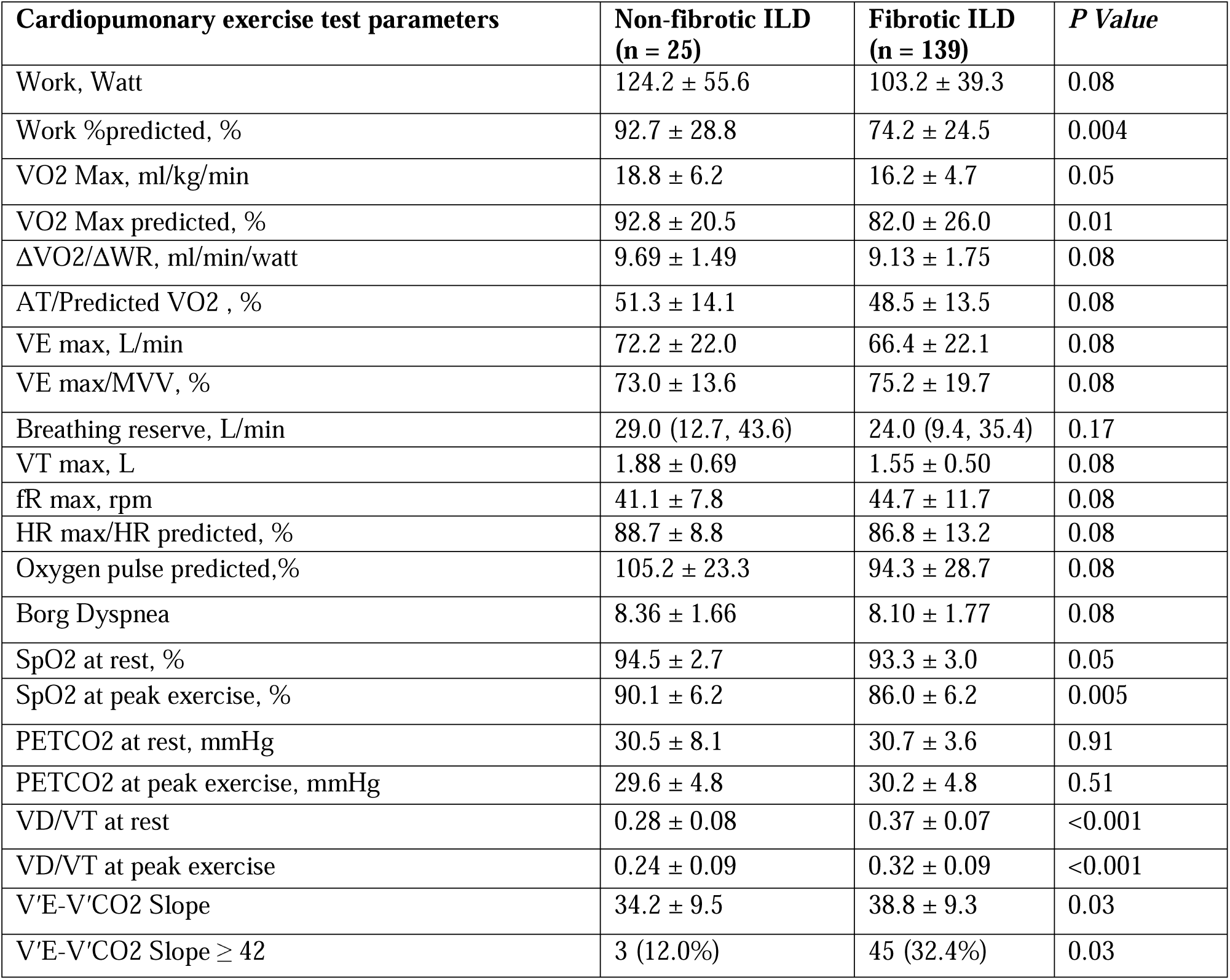
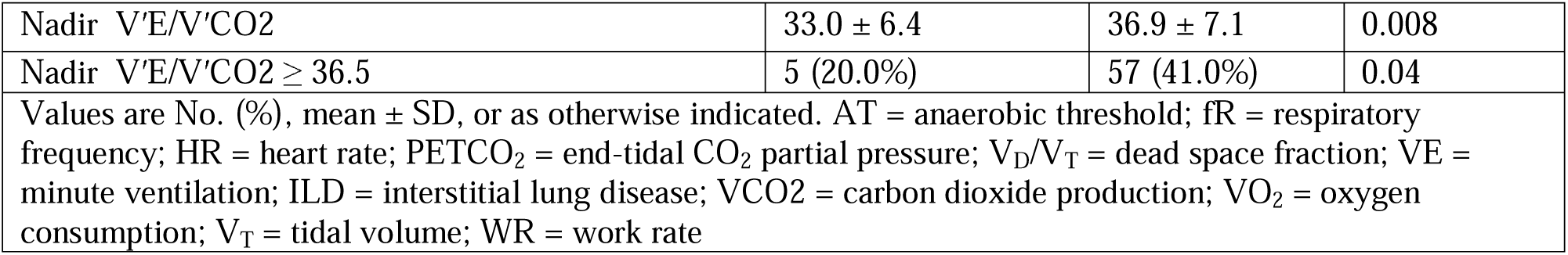
CPET variables in patients with non-fibrotic ILD (nonfibrotic HP) and fibrotic ILD (fibrotic HP and IPF).

**e-Table 2.** CPET variables in the overall study cohort categorized by DLCO.

|  | **Dlco by Z-score** |  |  |  |  |  |  |
| --- | --- | --- | --- | --- | --- | --- | --- |
| **Cardiopumunary exercise test parameters** | **Preserved (n = 73)** | **Mild (n = 31)** | **\*P Value** | **Moderate (n = 44)** | **\*P Value** | **Severe (n = 11)** | **\*P Value** |
| Work, Watt | 120.2 ± 44.4 | 107.0 ± 32.8 | 0.74 | 95.2 ± 37.7 | <0.001 | 77.5 ± 41.9 | <0.001 |
| Work %predicted, % | 88.2 ± 26.7 | 75.9 ± 20.6 | 0.09 | 67.4 ± 21.4 | <0.001 | 54.7 ± 13.4 | <0.001 |
| VO<sub>2</sub> Max, ml/kg/min | 18.9 ± 5.4 | 15.9 ± 3.6 | 0.01 | 14.7 ± 3.6 | <0.001 | 12.5 ± 4.7 | <0.001 |
| VO<sub>2</sub> Max predicted, % | 94.8 ± 26.5 | 82.1 ± 22.3 | 0.06 | 72.7 ± 18.4 | <0.001 | 63.5 ± 18.1 | <0.001 |
| $\Delta VO_2/\Delta WR$, ml/min/watt | 9.63 ± 1.66 | 9.20 ± 1.84 | 1.00 | 8.88 ± 1.45 | 0.10 | 7.76 ± 1.88 | <0.001 |
| AT/Predicted VO<sub>2</sub>, % | 52.9 ± 14.7 | 49.5 ± 11.8 | 1.00 | 44.5 ± 11.0 | 0.005 | 42.9 ± 13.4 | 0.11 |
| VE max, L/min | 72.1 ± 22.6 | 65.5 ± 21.7 | 0.94 | 65.5 ± 22.0 | 0.67 | 55.2 ± 12.5 | 0.10 |
| VE max/MVV, % | 75.9 ± 17.9 | 73.0 ± 20.1 | 1.00 | 75.7 ± 18.4 | 1.00 | 75.3 ± 25.9 | 1.00 |
| Breathing reserve, L/min | 20.9<br>(6.6 – 39.9) | 19.5<br>(12.1 – 39.6) | 1.00 | 22.0<br>(12.6 – 31.4) | 1.00 | 23.4<br>(12.4 – 37.6) | 1.00 |
| V<sub>T</sub> max, L | 1.71 ± 0.56 | 1.60 ± 0.43 | 1.00 | 1.52 ± 0.50 | 0.39 | 1.20 ± 0.29 | 0.01 |
| fR max, rpm | 43.8 ± 10.7 | 43.6 ± 10.8 | 1.00 | 44.4 ± 13.2 | 1.00 | 47.5 ± 9.8 | 1.00 |
| HR max/HR predicted, % | 89.6 ± 12.1 | 88.5 ± 10.2 | 1.00 | 84.3 ± 14.4 | 0.17 | 82.5 ± 11.7 | 0.49 |
| Oxygen pulse predicted,% | 107.2 ± 28.1 | 93.4 ± 26.2 | 0.09 | 84.4 ± 24.3 | <0.001 | 77.3 ± 20.6 | 0.003 |
| Borg Dyspnea | 8.03 ± 1.96 | 8.65 ± 1.08 | 0.63 | 7.93 ± 1.80 | 1.00 | 8.09 ± 1.81 | 1.00 |
| SpO<sub>2</sub> at rest, % | 94.1 ± 2.9 | 93.6 ± 2.2 | 0.86 | 93 .0 ± 3.2 | 0.21 | 90.5 ± 2.6 | 0.001 |
| SpO<sub>2</sub> at peak exercise, % | 90.3 ± 4.7 | 84.7 ± 5.7 | <0.001 | 83.8 ± 5.6 | <0.001 | 78.4 ± 5.0 | <0.001 |
| PETCO<sub>2</sub> at rest, mmHg | 30.8 ± 5.1 | 31.9 ± 4.0 | 0.69 | 29.8 ± 4.0 | 0.58 | 30.5 ± 3.2 | 0.99 |
| PETCO<sub>2</sub> at peak exercise, mmHg | 30.9 ± 3.8 | 30.7 ± 5.7 | 0.99 | 28.9 ± 5.5 | 0.12 | 28.5 ± 4.0 | 0.39 |
| V<sub>D</sub>/V<sub>T</sub> at rest | 0.34 ± 0.08 | 0.35 ± 0.07 | 0.98 | 0.40 ± 0.06 | <0.001 | 0.45 ± 0.08 | <0.001 |
| V<sub>D</sub>/V<sub>T</sub> at peak exercise | 0.26 ± 0.08 | 0.29 ± 0.07 | 0.55 | 0.36 ± 0.09 | <0.001 | 0.42 ± 0.07 | <0.001 |
| VE/VCO<sub>2</sub> Slope | 34.2 ± 5.0 | 37.5 ± 8.8 | 0.26 | 42.4 ± 11.9 | <0.001 | 46.4 ± 8.4 | <0.001 |
| VE/V'CO<sub>2</sub> Slope ≥ 42 | 7 (9.6%) | 10 (32.3%) | <0.001 | 19 (43.2%) | <0.001 | 9 (81.8%) | <0.001 |
| Nadir VE/VCO<sub>2</sub> | 33.5 ± 4.2 | 34.7 ± 5.1 | 0.78 | 39.6 ± 8.7 | <0.001 | 45.1 ± 9.5 | <0.001 |
| Nadir VE/VCO<sub>2</sub> ≥ 36.5 | 19 (26.0%) | 7 (22.6%) | 0.71 | 24 (54.5%) | 0.001 | 9 (81.8%) | <0.001 |

Values are No. (%), mean ± SD, or as otherwise indicated. AT = anaerobic threshold; fR = respiratory frequency; HR = heart rate; PETCO<sub>2</sub> = end-tidal CO<sub>2</sub> partial pressure; V<sub>D</sub>/V<sub>T</sub> = dead space fraction; VE = minute ventilation; VCO<sub>2</sub> = carbon dioxide production; VO<sub>2</sub> = oxygen consumption; V<sub>T</sub> = tidal volume; WR = work rate
\* *P* value, when compared to preserved pulmonary function based on Z score ≥ -1.65

**e-Table 3.**
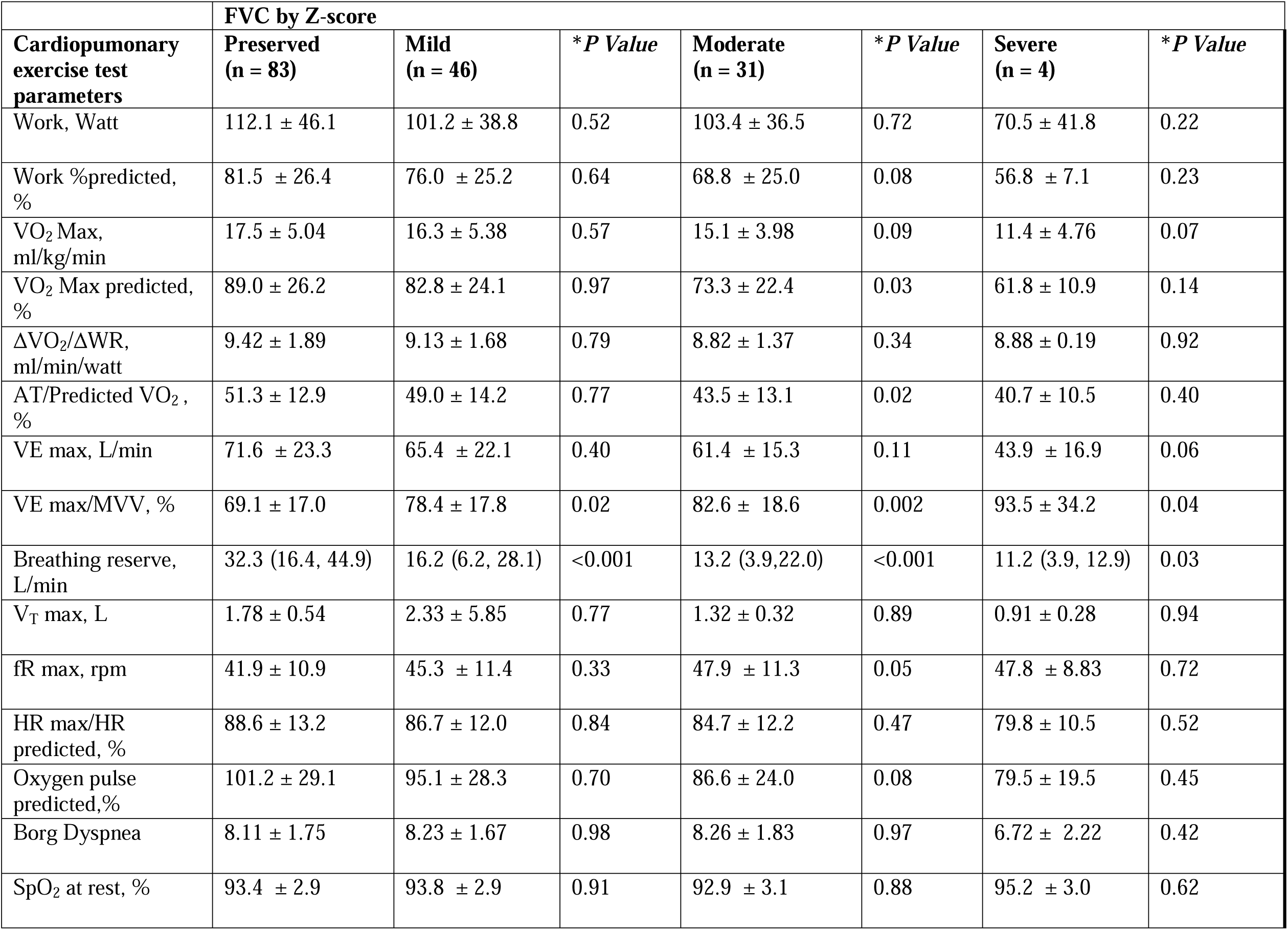
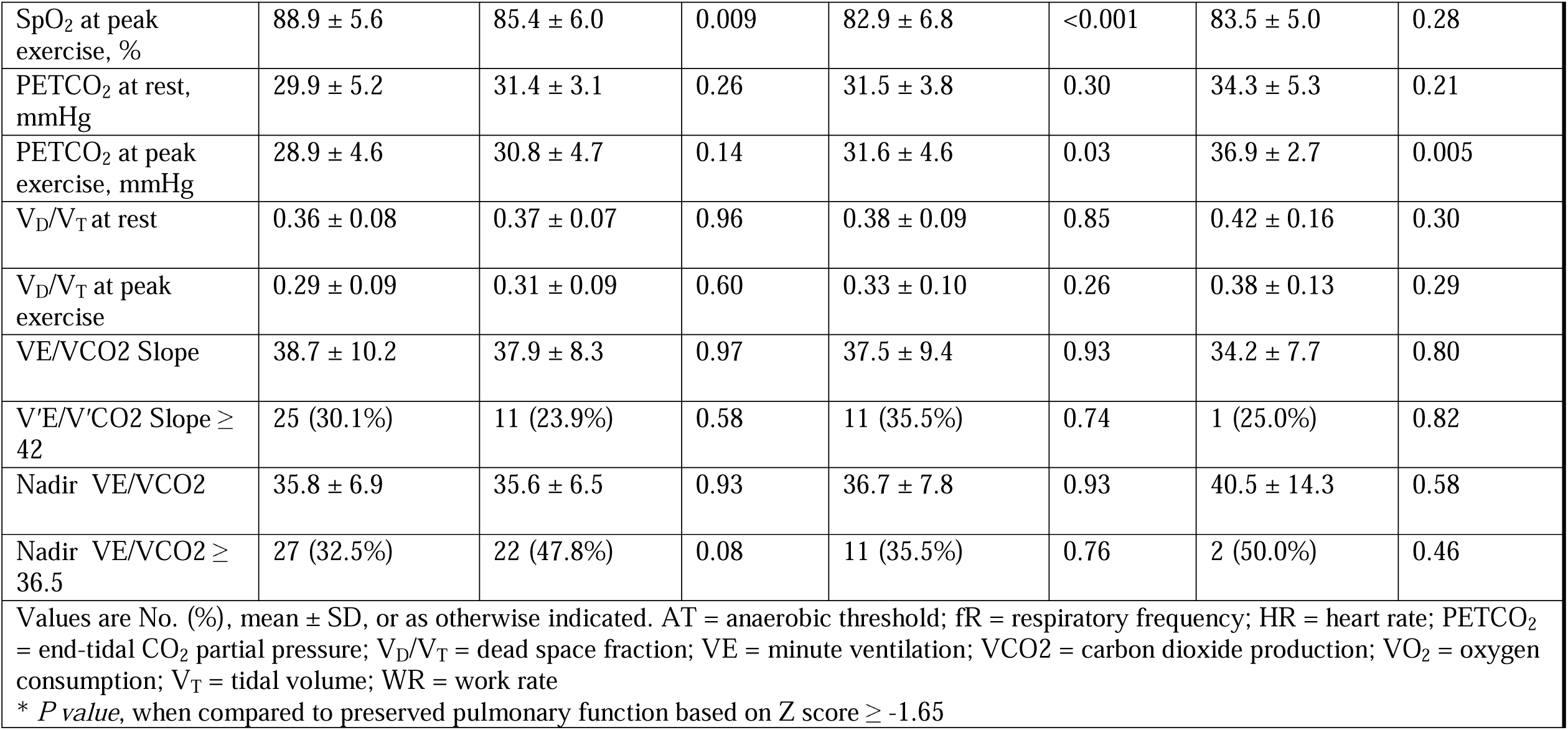
CPET variables in the overall study cohort categorized by FVC.

**e-Table 4.** CPET variables in the overall study cohort with and without pulmonary hypertension.

| **Cardiopulmonary exercise test parameters** | **No PH**<br>**(n = 107)** | **PH**<br>**(n = 57)** | ***P Value*** |
| --- | --- | --- | --- |
| Work, Watt | 112.2 ± 43.5 | 94.7 ± 38.8 | 0.008 |
| Work %predicted, % | 83.6 ± 26.9 | 64.5 ± 18.8 | <0.001 |
| VO<sub>2</sub> Max, ml/kg/min | 17.5 ± 5.1 | 14.9 ± 4.5 | <0.001 |
| VO<sub>2</sub> Max predicted, % | 89.0 ± 26.4 | 73.6 ± 20.3 | <0.001 |
| ΔVO<sub>2</sub>/ΔWR, ml/min/watt | 9.24 ± 1.80 | 9.16 ± 1.58 | 0.77 |
| AT/Predicted VO<sub>2</sub>, % | 50.1 ± 13.7 | 46.7 ± 13.1 | 0.11 |
| VE max, L/min | 69.4 ± 21.8 | 63.3 ± 22.2 | 0.09 |
| VE max/MVV, % | 73.9 ± 18.0 | 76.8 ± 20.5 | 0.36 |
| Breathing reserve, L/min | 23.4 (10.8, 40.6) | 18.3 (10.5, 29.7) | 0.07 |
| V<sub>T</sub> max, L | 1.66 ± 0.54 | 1.46 ± 0.47 | 0.01 |
| fR max, rpm | 43.3 ± 10.7 | 45.8 ± 12.0 | 0.18 |
| HR max/HR predicted, % | 88.9 ± 13.5 | 83.8 ± 10.3 | 0.007 |
| Oxygen pulse predicted,% | 101.2 ± 27.2 | 87.2 ± 28.1 | 0.004 |
| Borg Dyspnea | 8.04 ± 1.73 | 8.33 ± 1.79 | 0.30 |
| SpO<sub>2</sub> at rest, % | 93.8 ± 2.6 | 92.7 ± 3.4 | 0.03 |
| SpO<sub>2</sub> at peak exercise, % | 87.3 ± 6.3 | 85.4 ± 6.2 | 0.06 |
| PETCO<sub>2</sub> at rest, mmHg | 31.0 ± 4.8 | 30. ± 3.8 | 0.20 |
| PETCO<sub>2</sub> at peak exercise, mmHg | 30.4 ± 4.9 | 29.7 ± 4.7 | 0.43 |
| V<sub>D</sub>/V<sub>T</sub> at rest | 0.35 ± 0.08 | 0.39 ± 0.08 | 0.004 |
| V<sub>D</sub>/V<sub>T</sub> at peak exercise | 0.29 ± 0.09 | 0.35 ± 0.09 | <0.001 |
| V'E-V'CO<sub>2</sub> Slope | 37.1 ± 9.2 | 40.1 ± 9.6 | 0.05 |
| V'E-V'CO<sub>2</sub> Slope ≥ 42 | 29 (27.1%) | 19 (33.3%) | 0.40 |
| Nadir V'E/V'CO<sub>2</sub> | 35.0 ± 6.9 | 38.8 ± 6.9 | <0.001 |
| Nadir V'E/V'CO<sub>2</sub> ≥ 36.5 | 31 (29.0%) | 31 (54.4%) | <0.001 |
| Values are No. (%), mean ± SD, or as otherwise indicated. Values are No. (%), mean ± SD, or as otherwise indicated. AT = anaerobic threshold; fR = respiratory frequency; HR = heart rate; PETCO<sub>2</sub> = end-tidal CO<sub>2</sub> partial pressure; PH = pulmonary hypertension; V<sub>D</sub>/V<sub>T</sub> = dead space fraction; VE = minute ventilation; VCO<sub>2</sub> = carbon dioxide production; VO<sub>2</sub> = oxygen consumption; V<sub>T</sub> = tidal volume; WR = work rate |  |  |  |

**e-Figure 1.**
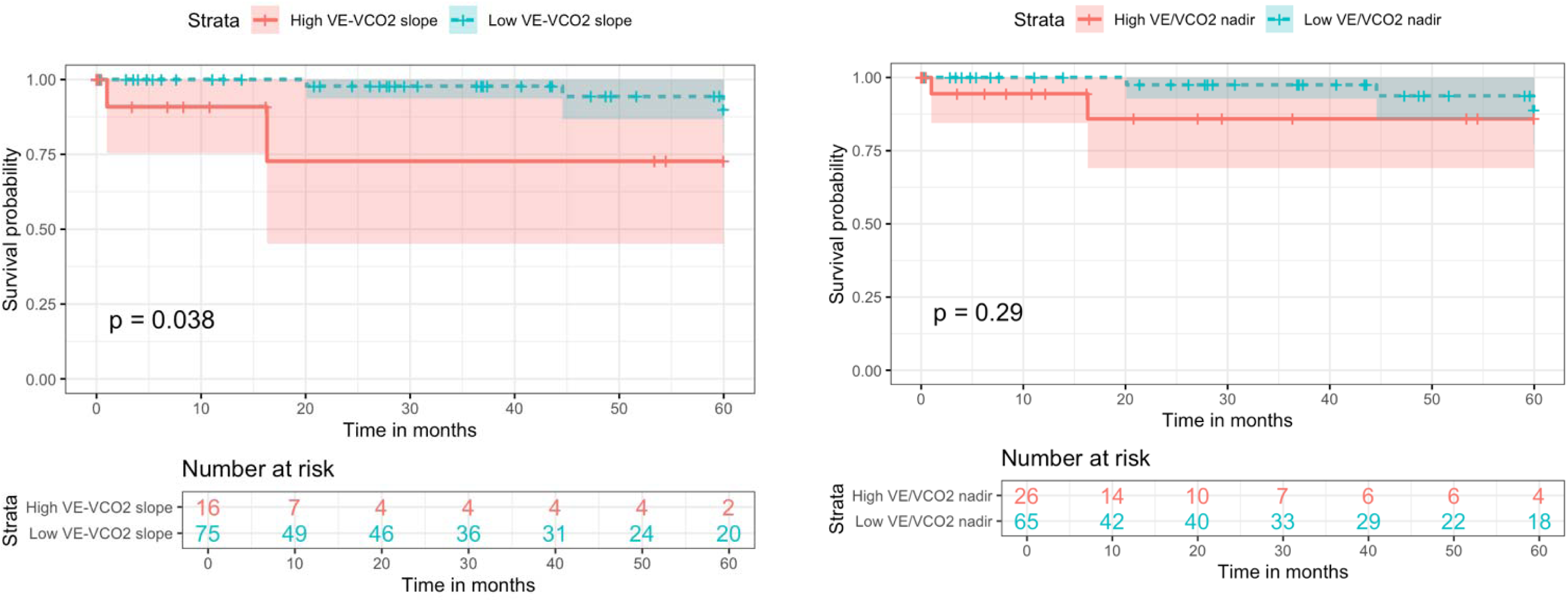
Assessment of Ventilatory Efficiency as a Predictor of 5-Year Survival in Patients with Hypersensitivity Pneumonitis: The VE/VCO2 slope is categorized as high (≥ 42) or low (< 42) (a), while the nadir VE/VCO2 is classified as high (≥ 36.5) or low (< 36.5) (b).

